# Early Imaging Identification of Osteoradionecrosis and Classification Using the Novel ClinRad System: Results from A Retrospective Observational Cohort

**DOI:** 10.1101/2025.01.29.25321211

**Authors:** MD Anderson Head and Neck Cancer Symptom Working Group; Collaborators, Jillian Rigert, Zaphanlene Kaffey, Zayne Belal, Lavanya Tripuraneni, Laia Humbert-Vidan, Ariana Sahli, Serageldin Attia, Katherine A. Hutcheson, Erin Watson, Andrew Hope, Cem Dede, Sudarat Kiat-amnuay, Muhammad Walji, Abdallah S.R. Mohamed, Vlad C. Sandulache, Clifton David Fuller, Stephen Y. Lai, Amy Moreno

**Author notes:** Co-First Authors (Jillian Rigert, Zaphanlene Kaffey, Zayne Belal).

## Abstract

**Objective:** Osteoradionecrosis of the jaw (ORNJ) is a chronic radiation-associated toxicity that lacks standardized classification criteria and treatment guidelines. Understanding early signs of tissue injury could help us better predict, prevent, and conservatively manage ORN. Our primary aims were to identify initial clinically-detected signs of ORN, determine the frequency of imaging-detected ORNJ, and validate the ability to classify cases using the novel system, ClinRad.

**Study Design:** A retrospective electronic health record review of 91 patients treated for head and neck cancer at The University of Texas MD Anderson Cancer Center with suspected ORN was performed by an Oral Medicine specialist to identify initial signs of ORN. Patients who received reirradiation to the head and neck or did not have enough evidence of ORN were excluded. A descriptive analysis was performed.

**Results:** 51 patients met the inclusion criteria. Half (53%) presented with imaging findings and exposed bone. Imaging findings in the absence of bone exposure were identified in 37%, of which disease progression was observed in 26%. All cases were classifiable using ClinRad.

**Conclusion:** Subclinical signs of bony changes consistent with ORN may be evident on imaging without exposed bone, supporting the use of imaging surveillance. ClinRad provided a mechanism to classify all cases at early onset.

**Data availability statement:** Anonymized data for the reported analyses is made publicly available on figshare at 10.6084/m9.figshare.28292186.

**Reporting guideline compliance statement:** In accordance with the EQUATOR Network (Enhancing the QUAlity and Transparency Of health Research) guidance, we have utilized the RECORD checklist, a guideline for the “REporting of studies Conducted using Observational Routinely-collected health data” (Benchimool El at al., 2015) The RECORD checklist is provided as a Supplementary file and available via 10.6084/m9.figshare.28292219.

Data was anonymized in accordance with the EQUATOR guideline “Preparing raw clinical data for publication: guidance for journal editors, authors, and peer reviewers” (Hrynaszkiewicz I et al., 2010).

## Introduction

Osteoradionecrosis (ORN) is a chronic radiation-associated toxicity that may become debilitating when progressive. Accurate identification and reporting of ORN among patients with head and neck cancer is complicated by a variety of presently used classification systems that differ in diagnostic criteria. The lack of standardization contributes to the wide range of cited prevalence rates, noted to be between 0.4% - 56% (Chronopoulos et al., 2018) and reduces the capacity to utilize aggregate datasets for analyses. There is also limited consensus on how to stage, grade, or manage ORN, including whether the use of Hyperbaric Oxygen (HBO) (Chuang, 2012; Forner et al., 2022) or pentoxifylline/vitamin E (Kolokythas et al., 2019) represent effective strategies. The ideal treatment strategy for optimal patient quality of life and wellness with lowest cost is the prevention of ORN through measures which include minimizing radiation toxicity, maximizing oral health care, routine screenings for early detection of potential risk factors, and early intervention when conservative treatment may stabilize or reverse disease process (Peterson et al, 2024). Even conservative measures for ORN may significantly increase financial burden on patients who often face financial toxicity and higher risk for psychosocial distress in the setting of head and neck cancer (HNC) and associated treatment. A 2019 report estimated conservative treatment for ORN may range from $4,000-$35,000 with some closer to $74,000 (Elting LS & Chang YC, 2019). HBO therapy would add significant time investment and financial burden on top of these reported costs with questionable efficacy for ORN management. Thus, prevention is essential, particularly when considering the financial and psychosocial toll that patients with head and neck cancer already face without the development of ORN, including increased risk for suicide secondary to financial and side effect burdens (Peterson et al., 2024).

To better understand the ORN disease process and efficacy of treatment strategies, consensus on the definition and classification system is necessary. Efforts by international interdisciplinary teams to create consensus guidelines for the classification and treatment of ORN have recently supported the use of the novel ClinRad system which considers both imaging and clinical evidence of disease independently (Table 1b) (Watson et al, 2024; Peterson et al, 2024). In contrast to the ClinRad classification criteria, several proposed definitions of ORN historically required clinical evidence of non-healing bone exposure (i.e., Marx, Epstein, Widmark) (Chronopoulous et al., 2018); however, we have shown that early bone physiologic changes due to radiotherapy (RT) can be detected sub-clinically/pre-symptomatically *before* clinical bone exposure or mucosal defect events (Joint Head and Neck Radiation Therapy-MRI Development Cooperative, 2020). In addition to 2-D panoramic images traditionally utilized in the dental setting for the monitoring of pathologic changes (orthopantograms), evidence supports the superiority of volumetric imaging including cone beam computed tomography (CBCT) (Gaêta-Araujo et al., 2021), computed tomography (CT) or magnetic resonance imaging (MRI) (Mallya & Tetradis, 2018; Miyamoto et al., 2021) in detecting early bony changes. Advantages of using CT or MRI include improved sensitivity, the ability to delineate areas of the affected bone, and the capacity to objectively quantify response to conservative medical interventions (i.e., pentoxifylline/vitamin E) (Head and Neck Radiation Therapy-MRI Development Cooperative, 2020).

Moreover, CT imaging features (radiomics) have similarly been shown to precede ORN development (Barua et al., 2021, Kamel et al., 2024), suggesting cortical alteration measured by CT attenuation may occur as a prodromal harbinger of clinical symptoms. The use of CT and MRI, which are captured serially before, during, and after radiation treatment for head and neck cancer surveillance, provides opportunities to acquire imaging biomarkers (i.e., dynamic contrast-enhanced magnetic resonance imaging (DCE-MRI) (Abdallah et al., 2020)) or structural metrics (i.e., Ultrashort TE (UTE) (Tyler et al., 2007) and Black Bone (van Dijk et al., 2020) sequences) to evaluate bony injuries correlated with subsequent risk of ORN development at early signs of disease development (Deshpande et al., 2015; Joint Head and Neck Radiation Therapy-MRI Development Cooperative, 2020). Moreover, just as the pathognomonic criteria defining ORNJ have been ambiguous, the lack of uniform disease designation within medical records due to the absence of a reliable ORNJ-specific International Classification of Diseases (ICD) code until the 2023 ICD-11 has hampered EHR data extraction as well as hampered coordinated efforts to aggregate data across institutes.

To our knowledge, assessment of the relative prevalence of “imaging only” ORN either as a preceding sign, or as a synchronous presenting sign with clinically exposed bone has not been previously explored. Thus, through a retrospective chart review, the authors aimed to characterize the initial signs of ORN radiographically and clinically and identify the temporal relationship between bony changes presenting on imaging and clinically as exposed bone. The authors classified ORN using the novel ClinRad system to assess the feasibility and capacity of said classification to capture ORN at observed disease severity at presentation, with secondary assessment of which (if any) diagnosis-related ICD designations was assigned by providers.

## Materials and Methods

### Patient Selection

A total of 1576 electronic records from a prospective single-center observational cohort of sequential head and neck cancer patients who underwent either definitive or adjuvant RT were retrieved and subsequently screened for the presence of ORN utilizing the institutional Standard Operating Procedure PA14-0947 “Osteoradionecrosis (ORN) Chart Abstraction.” The charts were searched using keywords correlating with Tsai’s classification of ORN (Table 1a) (Tsai et al., 2013). A secondary review was performed by an Oral Medicine specialist (OM) (JMR) using a manual search of keywords “osteoradionecrosis,” “osteonecrosis,” “ORN,” “exposed bone,” “bone exposure,” “fracture” to identify clinical and radiology notes with keywords suggestive of the presence of ORN. The cases were then (re)classified using the ClinRad grading system, (Table 1b) (Watson et al, 2024) with examples of each stage/grade shown in figure 1.

**Figure 1.**
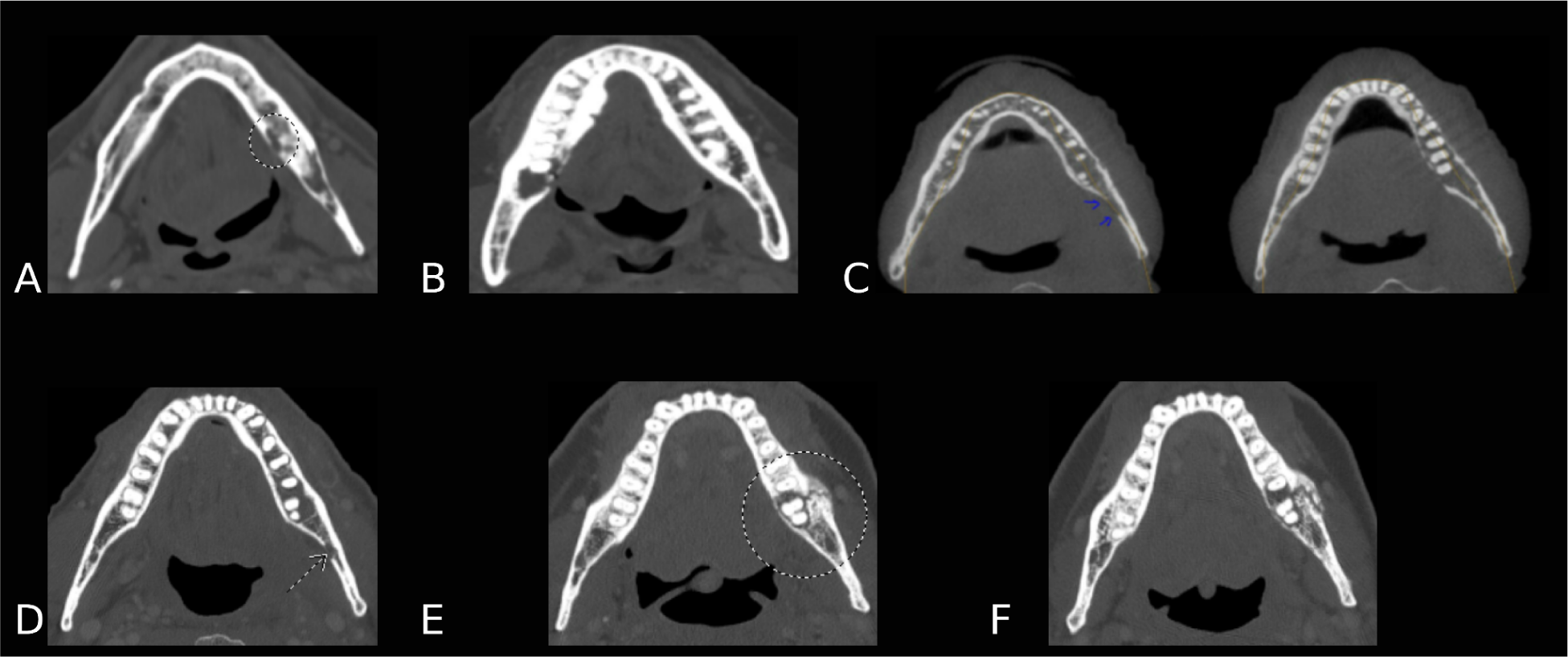
CT images representing the 5 Watson Stage/Grades: A.) Stage 0/Grade 1, B.) Stage 1/Grade 2, C.) Stage 2/Grade 3, D.) Stage 2/Grade 3, E.) Stage 3/Grade 4 at initial ORN diagnosis, F.) Stage 3/Grade 4 at four months with integral progression from initial ORN diagnosis.

**Table 1a.** The Tsai Grading System for Mandibular ORN.

| Grade | Definition | Search Terms |
| --- | --- | --- |
| 0 | No ORN | Discussed, potential side effects |
| 1 | Suspected on imaging, asymptomatic with surveillance, or minimal bone exposure with conservative management | Pentoxifylline/Trental, Tocopherol/Vitamin E, Antibiotics<br>Suspected ORN, Asymptomatic |
| 2 | Minor debridement received | Debridement<br>Sequestrectomy |
| 3 | Hyperbaric oxygen needed | HBO, Hyperbaric oxygen |
| 4 | Major surgery required (i.e., mandibulectomy) | Mandibulectomy, Mandible Reconstruction |

**Table 1b.** Watson et al, ClinRad ORN Classification System. ***(Derived from Watson et al, 2024)*.**

| Stage | Clinical Trial Grade | Radiographic Signs | Clinical Signs |
| --- | --- | --- | --- |
| Minor Bone Spicules<br>(Distinct from ORN) | 0 | None; Bony sequestra | Superficial mobile bone spicules or sequestra |
| 0 | 1 | Area of bone necrosis is confined to the alveolar bone | Mucosa intact |
| 1 | 2 | None or per Stage 0 | Exposed bone; includes pinpoint fistula and ability to probe to bone |
| 2 | 3 | Area of necrosis involves basilar bone or maxillary sinus | Mucosa intact or exposed bone |
| 3 | 4 | Pathologic fracture; Orocutaneous fistula; Oral antral communication/oral nasal communication | Pathologic fracture; Orocutaneous fistula; Oral antral communication/oral nasal communication |

A OM specialist manually reviewed images and diagnostic findings documented in clinician examinations and radiologist reports. For patients with reported ORN based on clinical and/or radiology reports, the OM completed an independent review of imaging features across all prior images/scans, in order to ascertain the earliest detectable sign of ORN, including the presence or absence of imaging-demonstrated alteration of bone either preceding or contemporaneous with the detection of ORN by clinical examination, and to verify the ability to classify ORN using the ClinRad system. Secondary aims were to identify the time-to-event (time from the end of RT to the initial signs of ORN) for patients treated with one course of RT to the head and neck with curative intent and to assess the rate of disease progression for patients who initially presented with imaging-only features. IRB waiver of informed consent for secondary imaging analysis was acquired under MDACC Protocol RCR 03-0800.

### Data Abstraction and Analysis

Demographics, primary tumor site, HPV/p16 status, and chemoradiation treatment received were recorded. Data from relevant notes, images, and problem lists were manually extracted using search terms as described above. Initial documentation of ORN diagnosis was identified (if present) and the clinical objective findings and imaging findings at the time of initial diagnosis were recorded. If ORN was identified on imaging, a manual search for the most proximal dental encounter was completed by filtering notes by “Enc Dept” (Encounter Department), “Main Dental.” The dental note was manually reviewed for objective findings, including whether the exposed bone was present or absent. Proximal clinical notes from medical professionals were also similarly reviewed. If the initial note of ORN was found clinically (by dental or medical clinicians), the reports from the most proximal computed tomography (CT), magnetic resonance imaging (MRI), cone beam CT (CBCT) and/or panoramic x-ray were evaluated for findings suggestive of ORN (i.e., cortical thinning, lytic lesions in the absence of recurrence, pathologic jaw fracture (Mallya and Tetradis, 2018).

Patients were excluded from further analysis if there were questionable/no signs of ORN (i.e., diagnosis confounded by primary dental disease process or dentigerous cyst, presence of metastatic disease in the area, or the absence of ORN features on imaging or clinically). Patients who received reirradiation to overlapping sites in the head and neck were also excluded to maintain clear time-to-event data following one course of radiation therapy (RT) with curative intent. For patients meeting inclusion criteria, ORN cases were separated into three groups based on the ORN characteristics identified at the initial presentation: imaging-only, imaging and clinical diagnosis, or clinical diagnosis only. The time from the end of RT to the initial signs of ORN was documented. For ORN cases that presented initially on CT only, the follow-up interval and state of disease progression from the time of initial presentation on CT to the most recent follow-up were recorded. All cases were classified by Stage/Grade using ClinRad.

To characterize our patient population, patient demographics, primary tumor characteristics and treatment received were evaluated in table 2. ORN at initial diagnosis was analyzed in Table 3 reporting number of patients, time to event, and Watson stage/grade across imaging-only, clinical-only, and combined cohorts. Additionally, we evaluated the total percentage for each Watson stage/grade by combining all subgroups. Finally, to identify common patterns of progression in patients post ORN diagnosis, we documented the disease course from the patient’s initial ORN diagnosis to their last follow-up visit.

**Table 2.** Demographics, Primary Tumor Characteristics, Treatment Received.

| **Sex** | **N (%)** |
| --- | --- |
| Male | 48 (94%) |
| Female | 3 (6%) |
| **Age (at time of ORN diagnosis)** | **Years** |
| Min | 43 |
| Max | 81 |
| Mean | 66 |
| **Deceased** | **N (%)** |
| Yes | 9 (18%) |
| No | 42 (82%) |
| **Primary Tumor Site** |  |
| Oropharynx (OPC) only | 49 (96%) |
| Both OPC/Oral cavity (OC) | 2 (4%) |
| **HPV/p16 status** |  |
| Positive | 45 (88%) |
| Negative | 4 (8%) |
| Unknown | 2 (4%) |
| **Radiation Treatment**<br>**Total Dose (Gy)** |  |
| Mean | 69 Gy |
| Min | 66 Gy |
| Max | 70 Gy |
| **Radiation Treatment**<br>**Fractions** |  |
| Mean | 33 |
| Min | 30 |
| Max | 35 |
| **Concurrent**<br>**Chemotherapy** |  |
| Yes | 49 (96%) |
| No | 2 (4%) |

**Table 3.** Characteristics of ORN at Initial Diagnosis.

| **Characteristics**<br>**(Imaging, Clinical,**<br>**Both)** | **# of Patients**<br>**(N = 51)** | **% Total**<br>**Patients**<br>**(N = 51)** | **TTE**<br>**(RTend to**<br>**ORNinitial)**<br>**Months** | **Watson**<br>**Stage(S)/**<br>**Grade (G)**<br>**N** | **Watson**<br>**(S)/(G)**<br>**%**<br>**of**<br>**SubGroup** |
| --- | --- | --- | --- | --- | --- |
| **Imaging\* Only**<br>**(Primarily CT)** | 19 | 37% | Mean :- 21 | S0/G1 :- 12 | 63% |
|  |  |  | Min :- 6 | S1/G2 :- 0 | 0% |
|  |  |  |  | S2/G3 :- 6 | 32% |
|  |  |  | Max :- 52 | S3/G4 :- 1 | 5% |
| **Clinical Only**<br>**(Exposed Bone or**<br>**Pain w/ ORN**<br>**Diagnosis)** | 5 | 10% | Mean :- 5 | S0/G1 :- 0 | 0% |
|  |  |  | Min :- 4 | S1/G2 :- 5 | 100% |
|  |  |  |  | S2/G3 :- 0 | 0% |
|  |  |  | Max :- 6 | S3/G4 :- 0 | 0% |
| **Imaging\* and**<br>**Clinical**<br>**(Primarily CT)** | 27 | 53% | Mean :- 21 | S0/G1 :- 0 | 0% |
|  |  |  | Min :- 4 | S1/G2 :- 23 | 85% |
|  |  |  |  | S2/G3 :- 3 | 11% |
|  |  |  | Max :- 62 | S3/G4 :- 1 | 4% |
| **Watson Stage**<br>**(S)/Grade (G)**<br>**Total**<br>**(ALL Subgroups**<br>**Combined)** | **N = 51** | **%** |  |  |  |
| S0/G1 | 12 | 24% | Mean :- 22 |  |  |
|  |  |  | Min :- 6 |  |  |
|  |  |  | Max :- 52 |  |  |
| | | | Median ± SD<br>: $19 \pm 14.40$ | | |
|  |  |  | 95% CI :<br>14.18 - 30.82 |  |  |
| S1/G2 | 28 | 55% | Mean :- 18 |  |  |
|  |  |  | Min :- 4 |  |  |
|  |  |  | Max :- 67 |  |  |
| | | | Median ± SD<br>: $10 \pm 18.64$ | | |
| | | | 95% CI ±<br>10.34 - 24.80 | | |
| S2/G3 | 9 | 18% | Mean :- 19 |  |  |
|  |  |  | Min :- 7 |  |  |
|  |  |  | Max :- 44 |  |  |
| | | | Median ± SD<br>: $17 \pm 12.04$ | | |
|  |  |  | 95% CI 9.86<br>- 28.36 |  |  |
| S3/G4 | 2 | 4% | Mean :- 34 |  |  |
|  |  |  | Min :- 24 |  |  |

### Data anonymization

Patient data was anonymized by removing all patient health information that could potentially risk identification of patients and a key was made available to internal authors. All dates were removed and replaced with time-to-event data. Ages are reported from time of initial ORN. Initial ORN is reported in months from the time between the end of RT to the first signs of ORN.

## Results

### Patient Selection

Of the 91 patients’ charts manually reviewed, 51 patients met all inclusion criteria and displayed clinical and/or radiographic signs of ORN. Of the 40 excluded patients, 24 (26%) had received prior RT to overlapping sites in the head and neck and 16 patients displayed no signs of ORN or signs of suspected primary odontogenic or oral infection without clear signs of coincident ORN development.

Of the 51 patients meeting inclusion criteria, 48 were male (94%) with a mean age of 66 years (range 43-81) at the time of initial signs of ORN. The primary tumor site was located in the oropharynx in 100% of these patients, with two patients having tumors that spanned the oral cavity and oropharynx regions, and 88% of tumors were HPV/p16 positive. The included patients received one course of head and neck RT with a mean radiation dose of 69 Gy (range 66-70 Gy) delivered over a mean of 33 fractions (range 30-35 fractions). Most patients (96%) received concurrent chemotherapy (Table 2).

### EHR ORN Data Collection ICD Documentation

Approximately 50% of included patients did not have an International Statistical Classification of Diseases and Related Health Problems (ICD) code for ORN documented in the Problem List at any timepoint. Of the patients who did have an associated ICD code designated in the Problem List, the majority (96%) were listed with ICD-10 M27.2, a non-specific code for “Inflammatory Conditions of Jaws.” One patient had M87.2 “Osteonecrosis due to previous trauma” and one patient was diagnosed with both M27.2 and M87.2 ICD codes.

### Initial Signs of ORN

Of the 51 patients confirmed to have features consistent with ORN, 19 (37%) patients displayed initial signs on imaging only *withou*t reporter clinical bone exposure (Table 3). Of these 19 patients, 5 (26%) had usbsequent imaging progression of ORN with 2 displaying interval progression on CT then stabilizing, 1 developing exposed bone following dental extraction then stabilizing and 2 needing a hemimandibulectomy for treatment of ORN. For the 2 patients requiring surgical intervention, one patient presented with a mandible fracture visible on CT, pain and paresthesia of his left lower lip without exposed bone as the initial sign of ORN and one developed a jaw fracture 12 months after the initial ORN signs on CT. The mean follow-up interval for the patients who presented with CT-only findings at the initial presentation of ORN was 32 months (Table 4). Two patients were excluded from the mean calculation as one patient did not have a follow-up scan and one patient was lost to follow-up at 3 months.

**Table 4.** Initial Presentation Imaging Only (N = 19) - Progression From Initial ORN to Last Follow-up.

| **Progression** | **N (%)** | **Notes** |
| --- | --- | --- |
| Stable | 10 (53%) |  |
| Progressed | 5 (26%) | 2 on CT only - interval progression then stabilized;<br>1 exposed bone following dental extraction then stabilized;<br>2 needing surgical intervention |
| Improved | 2 (11%) | Stable to improved (may be difference in imaging angle, contrast) |
| Unable to determine* | 2 (11%) | *No subsequent imaging (1) and lost to follow-up (1) |
| **Time from Initial ORN to last follow-up visit** | **Months** | *Two patients excluded from the below calculations due to having no subsequent imaging (1) and being lost to follow-up (1) |
| Mean | 32 |  |
| Min | 11 |  |
| Max | 51 |  |

### ORN Staging & Time-to-Event

Using the ClinRad ORN Classification System, 12 (24%) patients presented with Stage (S) 0/Grade (G) 1 ORN, 28 presented with S1/G2 (55%), 9 (18%) presented with S2/G3, and 2 (4%) presented with S3/G4 at the initial signs of ORN. The time from the end of RT to the initial signs of ORN (Time to Event (TTE)) is provided along with the initial signs (imaging only, clinical only, both) and Watson Stage/Grade in Table 3.

## Discussion

Patients with head and neck cancer treated with RT have a lifetime risk of developing ORN. Despite awareness of ORN complications since the 1920s, the definition and diagnostic criteria continue to be challenged by competing perspectives. Several classification systems for ORN rely on the presence of bone exposure which may delay the diagnosis. However, the use of imaging surveillance, such as the CT scans captured during cancer treatments, may provide a pathway to identify normal tissue changes that precede the development of clinically apparent ORN. This information may serve to improve our understanding of the disease as well as our ability to predict and prevent ORN development and progression, improve patients’ quality of life and reduce treatment costs.

In 2024, a joint guideline publication from the American Society of Clinical Oncology (ASCO) and the Multinational Association of Supportive Care in Cancer-International Society of Oral Oncology (MASCC-ISOO) aimed at providing guidance for the prevention, assessment, classification (i.e., grading) and management of osteoradionecrosis (ORN) of the jaws (Peterson et al., 2024). The expert panel established that ORN may be defined without depending on factors such as clinical signs/symptoms or a duration of time, which is consistent with the findings of this current study. The guideline endorsed the use of the ClinRad system which was developed using evidence-based support for the classification of ORN of the maxilla and mandible and meets criteria established by Shaw et al. (2017) for the classification of ORN in clinical trials. Further, ClinRad is compatible with interprofessional use across the dental and medical sectors which have varying forms of clinical and imaging assessment protocols. For these reasons, the authors utilized the ClinRad system to confirm its utility and capacity to capture early-stage disease.

All of the patients identified as having ORN were able to be classified into the ClinRad system ORN based on clinical notes, radiology reports, and imaging review by OM, whereas prior systems (including the internally-developed Tsai et al. classification) would have elided those with radiographic but non-visible/probable bone. Due to the subjective nature of ORN diagnosis and lack of histologic confirmation, validation via interrater confirmation across ClinRad (and/or other scales) may be performed to strengthen the confidence in the diagnosis and assigned staging/grading. Select cases with more ambiguous ORN features were coreviewed by a head and neck radiation oncologist (ACM) for verification of the presence of suspected ORN and ClinRad Stage/Grade assigned independent of OM Stage/Grade to avoid bias.

In alignment with the ClinRad system, the diagnosis of ORN for this retrospective study was considered when evident on imaging alone, when clinical and radiographic signs were present, and when clinical signs/symptoms were present in the absence of reported changes on CT, MRI, CBCT or panoramic x-ray. A study strength is that dental, medical, and imaging reports were available due to our institution’s integrated electronic health record system which enhanced data completeness. The imaging was also available for personal review by the OM to verify and classify the stage/grade of ORN if present, reducing reliance on radiology reports alone and enabling the ability to assess the precise location of bony changes. This study examined the Stage/Grade of ORN at initial presentation and the progression for cases that initially presented on imaging only; future research to be explored includes monitoring all cases over time, including which interventions were provided, to explore features of disease present at various time-points to better understand and define the disease process as well as the efficacy of interventions.

A limitation of this study, and ORN studies in general, is that the presence of ORN was not confirmed histologically as biopsy confirmation for ORN is typically not indicated as it may lead to disease progression in an at-risk, irradiated field. This leads to reliance on subjective assessments and lower confidence in the ORN diagnosis versus another disease process. For example, the diagnosis of ORN on imaging was confounded by the presence of dental disease and infections which may appear with similar lytic changes in the bone. Additionally, these diseases may occur together as patients with dental disease and infections are more susceptible to developing ORN (Kojima et al., 2022). Of the 91 originally included cases, 16 cases were excluded due to insufficient findings consistent with ORN and/or a primary presentation of dental disease or infection without differentiating features consistent with ORN.

For patients who presented with clinical signs of ORN without bony changes on the CT, it is possible that bony changes were not detected due to their often subtle nature or the areas were obscured by dental artifacts. The exposed bone observed clinically may also represent a mucosal breakdown without apparent bony changes which may put into question whether the process truly represents ORN.

Due to the lack of standardized diagnostic criteria and recording in the EHR, the data collected for this study was largely unstructured and required manual data extraction which was time-consuming and prone to human error. At the time of this study, ICD-10 was in use which does not have a specific code for ORN (this has been addressed in the ICD-11 revision (Peterson et al., 2024)). The lack of concrete diagnostic criteria and ICD-10 code specific for ORN likely contributed to 50% of the patients not having ORN coded in the Problem List. If patients would have been excluded from initial screening using structured data such as the diagnosis of ORN on the Problem List, nearly half of the included patients would have been omitted.

Further research should be aimed at the implementation of a standard classification system of ORN (such as ClinRad), the development of consensus-based/validated diagnostic criteria and nomenclature (Peterson et al., 2024; International Oral Consortium, 2024 pre-print), and understanding the utility of CT and/or MRI imaging for monitoring of normal tissue changes, including vasculature and bone, which may predict and detect early changes consistent with ORN. Standard evidence-based treatment guidelines for conservative treatment of ORN also remain unmet needs and warrant further exploration. Prevention of ORN remains the strongest form of intervention, and continued efforts to optimize medical and dental health along with reducing radiation toxicity to normal tissues remain a priority.

### Conclusions

This study revealed that early signs of bony changes consistent with ORN development may be evident on imaging without clinical signs of exposed bone, with disease progression in over a quarter of the cases initially presenting with imaging-only ORN. The inclusion of imaging-only features in the definition of ORN (as is present in the ClinRad system) and use of imaging-based surveillance protocols (in addition to clinical examination) are recommended. All ORN cases were able to be classified using the ClinRad system, validating the inclusivity of the system albeit the clinical utility warrants further investigation. To improve the prevention, surveillance, diagnosis and treatment of ORN, there is a high need for evidence-based, multidisciplinary consensus on the definition of ORN, surveillance recommendations, treatment, and reporting methods for patients with HNC treated with RT.

## Declaration of Generative AI and AI-assisted technologies in the writing process

During the preparation of this work, we utilized ChatGPT (GPT-4 architecture) to enhance the grammatical accuracy and semantic structure of certain sections of the text. After using this tool/service, the author(s) reviewed and edited the content as needed and take(s) full responsibility for the content of the publication.

## Supplemental Materials

Data descriptor is available on figshare at DOI 10.6084/m9.figshare.28306514. <u>PA14-0947 “Osteoradionecrosis (ORN) Chart Abstraction”</u>

## Acknowledgments

This research was accomplished within The University of Texas MD Anderson Cancer Center-Oropharynx Cancer Program generously supported (in part) by Mr. & Mrs. Charles W. Stiefel.

